# Detecting schizophrenia, bipolar disorder, psychosis vulnerability and major depressive disorder from 5 minutes of online-collected speech

**DOI:** 10.1101/2024.09.03.24313020

**Authors:** Julianna Olah, Win Lee Edwin Wong, Atta-ul Raheem Rana Chaudhry, Omar Mena, Sunny X. Tang

**Affiliations:** Psyrin Ltd. London, UK; Psychiatry Research, Feinstein Institutes for Medical Research, Department of Psychiatry, Donald and Barbara Zucker School of Medicine, Department of Psychiatry, Zucker Hillside Hospital

## Abstract

**Background:** Psychosis poses substantial social and healthcare burdens. The analysis of speech is a promising approach for the diagnosis and monitoring of psychosis, capturing symptoms like thought disorder and flattened affect. Recent advancements in Natural Language Processing (NLP) methodologies enable the automated extraction of informative speech features, which has been leveraged for early psychosis detection and assessment of symptomology. However, critical gaps persist, including the absence of standardized sample collection protocols, small sample sizes, and a lack of multi-illness classification, limiting clinical applicability. Our study aimed to (1) identify an optimal assessment approach for the online and remote collection of speech, in the context of assessing the psychosis spectrum and evaluate whether a fully automated, speech-based machine learning (ML) pipeline can discriminate among different conditions on the schizophrenia-bipolar spectrum (SSD-BD-SPE), help-seeking comparison subjects (MDD), and healthy controls (HC) at varying layers of analysis and diagnostic complexity.

**Methods:** We adopted online data collection methods to collect 20 minutes of speech and demographic information from individuals. Participants were categorized as “healthy” help-seekers (HC), having a schizophrenia-spectrum disorder (SSD), bipolar disorder (BD), major depressive disorder (MDD), or being on the psychosis spectrum with sub-clinical psychotic experiences (SPE). SPE status was determined based on self-reported clinical diagnosis and responses to the PHQ-8 and PQ-16 screening questionnaires, while other diagnoses were determined based on self-report from participants. Linguistic and paralinguistic features were extracted and ensemble learning algorithms (e.g., XGBoost) were used to train models. A 70%-30% train-test split and 30-fold cross-validation was used to validate the model performance.

**Results:** The final analysis sample included 1140 individuals and 22,650 minutes of speech. Using 5- minutes of speech, our model could discriminate between HC and those with a serious mental illness (SSD or BD) with 86% accuracy (AUC = 0.91, Recall = 0.7, Precision = 0.98). Furthermore, our model could discern among HC, SPE, BD and SSD groups with 86% accuracy (F1 macro = 0.855, Recall Macro = 0.86, Precision Macro = 0.86). Finally, in a 5-class discrimination task including individuals with MDD, our model had 76% accuracy (F1 macro = 0.757, Recall Macro = 0.758, Precision Macro = 0.766).

**Conclusion:** Our ML pipeline demonstrated disorder-specific learning, achieving excellent or good accuracy across several classification tasks. We demonstrated that the screening of mental disorders is possible via a fully automated, remote speech assessment pipeline. We tested our model on relatively high number conditions (5 classes) in the literature and in a stratified sample of psychosis spectrum, including HC, SPE, SSD and BD (4 classes). We tested our model on a large sample (N = 1150) and demonstrated best-in-class accuracy with remotely collected speech data in the psychosis spectrum, however, further clinical validation is needed to test the reliability of model performance.

## Introduction

Alterations of speech in psychiatric disorders has been proposed as an objective, reproducible, and time-efficient biomarker (1–3) given its high prognostic value for onset, course, chronicity, and treatment response (4–16) across numerous conditions. Objective and scalable speech markers have gained special interest in psychosis where they can be combined with machine learning (ML) algorithms to detect illness, gauge symptom severity or predict relapse from speech (13,16–21).The putative predictive power of speech for psychotic disorder prognosis and development of psychosis from at risk state has received particular attention, as this opens up the premise of early, objective detection and targeted prevention (9,10,21–23). Importantly, speech and language were directly linked to key symptoms of psychosis, including formal thought disorder, changes in vocal expression, and flattened affect (24–29). Several speech features, including pitch, rhythm, and connectivity could signal changes in motor control function and neural connectivity, which correlate with psychotic symptom severity (30–34). These connections provided a neurobiological and phenomenological basis for why speech-based assessment can be useful in psychosis.

Despite these advancements, several critical gaps remain in the existing literature, limiting the application of speech analysis and assessment in clinical settings. Firstly, there is no standardized speech sample collection procedure. Speech elicitation methods hugely vary across research, with varying lengths and protocols. Prior studies often used speech recordings from laboratory settings with intense human involvement in terms of recording and transcription which limits the comparability of findings. Secondly, partly driven by the burden of recruitment and assessment procedures, previous studies have been conducted with relatively limited sample sizes and relatively narrow clinical scope (e.g., patients with acute psychotic episodes). This limits the complexity of applicable computational modelling of speech data and does not address the translation of findings to more heterogenous clinical populations. Thirdly, research to date has focused on primarily binary classifications between “healthy” individuals and individuals with one specific disorder, like bipolar or schizophrenia. This does not translate to the discriminatory diagnosis or detection of subtle changes that clinicians perform routinely.

Moreover, the focus on binary discrimination provides limited insight into whether speech-based ML models can capture disorder-specific alteration in speech rather than learning the difference between healthy and unhealthy speech patterns.

To address these gaps, we collected speech data with various elicitation procedures remotely, online, from a large cross-diagnostic sample of individuals (N = 1140). Participants included those at different points on the schizophrenia-bipolar continuum - people who have been diagnosed with schizophrenia spectrum disorders (SSD), people who have been diagnosed with bipolar disorder (BD) and people who have sub-clinical psychotic experiences (SPE) suggesting some vulnerability to psychosis but without meeting the diagnostic criteria of full-blown psychotic conditions and healthy control subjects, who might experience some distress regarding their mental health but have not received any psychiatric diagnosis or experience SPE (HC). Based on significant biological, genetic, neurological and phenomenological overlap, the schizophrenia-bipolar continuum can be understood as spanning the psychosis spectrum, where one dimension represents the affective nature of the condition (from affective to non-affective psychosis) and another dimension represents the severity and clinical nature of the symptoms (from sub-clinical experiences to full-blown, clinical conditions) (30,35–38). Studies that aimed to identify biomarkers and intermediate phenotypes found that there is an unexpectedly high overlap in the biological characteristics of psychotic diseases across the schizophrenia and bipolar spectrum and clear distinctions based on biomarkers are not apparent which underlines the continuum interpretation (37,38). However, within the current diagnostic system and at current level of scientific understanding, it is still worthwhile to distinguish between BD and SSD as they have different prognoses and may require different treatments, therefore we wanted to test whether speech markers can capture these differences.

In addition to the primary clinical focus on the schizophrenia-bipolar spectrum, we collected speech data from people with major depressive disorder (MDD) to serve as a non-psychotic comparison sample. MDD is a highly prevalent condition which overlaps with psychosis both with regard to comorbid patients and shared clinical presentations (e.g., anhedonia, amotivation) (30,39–41). Also, MDD has been also connected to speech alterations that can overlap with speech changes observed in the psychosis spectrum, posing a potential challenge to speech-based discriminatory diagnosis (39,40). Lastly, we collected speech from healthy individuals (HC) to provide a control condition and help to evaluate the discriminatory power of speech further by adding additional layering to out analyses.

We created a ML pipeline adopting state-of-the-art text-based and paralinguistic approaches and tested its ability to perform classification tasks across multiple conditions. In brief, our study aimed to:

1. Identify an optimal assessment approach for the online and remote collection of speech, in the context of assessing the psychosis spectrum.
2. Evaluate whether our ML pipeline can discriminate among different conditions on the schizophrenia-bipolar spectrum (SSD-BD-SPE), help-seeking comparison subjects (MDD), and healthy controls (HC) at varying layers of analysis.

## Methods

### Participants

Participants from the United States of America or the United Kingdom were recruited via the online platform Prolific (https://www.prolific.co). Potential participants were screened via self-report for the inclusion and exclusion criteria (N = 10,000) Diagnostic group was determined based on self-reported prior clinical diagnosis and responses to the 16-item prodromal questionnaire(42,43) and the 8-item Patient Health Questionnaire (44–46). Inclusion criteria included age of 18 years or above, native speaker of English, capability to record voice on adigital device, and ability to provide informed consent. We labelled participants into the following groups:

1. **Healthy control (HC)**: no history of a diagnosed mental disorder **and** scored less than 3 on both the PQ-16 symptom scale (prodromal psychotic symptoms) and PHQ-8 (depression symptoms) (42,43,46)
2. **Sub-clinical psychotic experiences (SPE):** no history of any diagnosed psychiatric disorder **and** scored more than 6 on the PQ-16 symptom scale (prodromal psychotic symptoms), a proxy for clinical at-risk mental state (42).
3. **Bipolar disorder (BD):** reported clinical diagnosis of bipolar disorder (type I or II) but did **not** report diagnosis of schizophrenia or schizoaffective disorder. Participants who reported co-morbid conditions other than schizophrenia and schizoaffective disorder were included, e.g., generalized anxiety, substance use disorders. Notably, participants in BD group have not necessarily experienced psychotic symptoms.
4. **Schizophrenia-spectrum disorders (SSD):** reported clinical diagnosis of schizophrenia or schizoaffective disorder but did **not** report diagnosis of bipolar disorder. Participants who reported co-morbid conditions other than bipolar disorder were included, e.g., depression, anxiety.
5. **Major depressive disorder (MDD):** reported diagnosis of major depression **and** did **not** report a diagnosis of bipolar disorder, schizophrenia, schizoaffective disorder **and** scored less than 3 on the PQ-16. Participants who reported co-morbid conditions other than BD or SSD conditions were included.

Exclusion criteria included diagnosis with autism spectrum disorder or other developmental disorders which affect speech and language, history of cognitive impairment or dementia. Written, informed consent was collected from all participants.

### Online Assessment of Speech

Speech was collected from eligible participants using an online experiment platform where participants provided secure voice samples in response to standardised prompts. Participants were instructed to record in a quiet environment, alone, and explicitly asked to avoid conditions that can reduce the quality of the audio recording. After an initial microphone test, each participant underwent five different speech-based tasks (detailed in **Table 1**), taking between 16- 20 minutes to complete. We selected the five tasks based on use in previous research (17,22,47,48). Participants were allowed to stop and resume the testing procedure within a 1- week window.

**Table 1:** Description of different speech elicitation tasks.

| Speech tasks in the data collection protocol |  |  |
| --- | --- | --- |
| Name | Prompts | Time |
| Dream recall | Participants were asked to recall one of their recent dreams. | 1 min |
| Picture description | Participants were shown 8 different ambiguous, black-and-white images from the Thematic Apperception Test (TAT) and asked to describe each of them and how they made them feel for one minute each. | 8 mins |
| Free speech | Participants were asked to discuss 4 neutral topics ('How was your last weekend?', 'What do you like about your work?', 'Could you describe your favourite food?', 'How and where was your last holiday?' ), each for one minute. | 4 mins |
| Neutral text reading | Participants were asked to read a brief, prewritten, neutral text. The text was a short version of the fable, the "North Wind and the Sun" (full text in Appendix 1) | ~ 1 min |
| Emotional text reading | Participants were asked to read three brief, prewritten, emotional bedtime stories (angry, happy, scared) (full text in Appendix 2-5) | ~ 3 mins |

Participants were compensated for their time. For screening, participants were paid 3 GBP, and for the speech recording task, participants were paid 9 GBP, via Prolific.

### Data Pre-processing

Incoming data was uploaded to a secure Cloud server. Audio data was automatedly transcribed into text using Amazon Transcribe API. Files that contained less than 5 words/task were excluded from analyses, and stop and filler words (e.g. hm, ahmm, uhmm, hmm) were removed before analysing the text, using a predefined list of targeted words that suited our Amazon ASR system. Audio files that contained stereo WAV formats were converted to mono format.

### Feature Extraction

Following pre-processing, both raw audio data and text transcripts went through a feature extraction pipeline. Features were selected to complement each other and reflect abnormalities at different levels of language.

#### Text-based features

NLP techniques were applied to extract features of abnormal language across semantics, morphology and syntactic levels.

Semantic coherence was captured with 7 features. We first applied word-embedding methodology: each word was represented as a vector, such that words used in similar contexts (e.g. ‘desk’ and ‘table’) were represented by similar vectors. Vector representations were generated by using word embeddings from the pre-trained word2vec Google News model (49) . Based on word representations as vectors, sentence embedding was calculated using Smooth Inverse Frequency (SIF), to produce a single vector for each sentence (29).We used word2vec and SIF embeddings because they previously gave the greatest group differences between patients with schizophrenia and control subjects and have been widely applied in psychosis research (22,29,47,49,50). Finally, cosine similarities between adjacent sentences and words were calculated to represent coherence at several levels. Coherence features included mean coherence score of the sentences, mean coherence per every 10 tokens, changes in sentence coherence score over the text, repetition, repetition divided by number of tokens in the speech sample, mean sentence coherence divided by the number of tokens in the speech sample and mean number of tokens per sentences.

Speech connectivity was measured by 30 features. Speech connectivity captures both semantic and syntactic information, measuring both the semantic coherence, syntactic complexity, grammatical complexity and correctness of language use. Overall, connectivity measures are considered to provide a proxy of narrative planning(51) as they can determine markers of short-range and long-range recurrence of a subject in the text (52). To measure speech connectivity, we generated speech graphs from the text, where all of the words represented as nodes in a graph while the grammatical connections between words were represented as edges in the graph, following the procedure of Mota et al(51). Graphs were generated using three different types of text pre-processing, and different parameters of the graphs (e.g., the largest connected component, number of repeated edges, diameter) were used as connectivity features, to capture the relation of syntactic and semantic information.

Syntactic complexity was measured by 6 features. These metrics were based on automated part-of-speech tagging using NLTK (Natural Language Toolkit) Python package. Our features measured the frequency of tags compared to a number of tokens and the frequency of tags compared to each other (e.g., Number of Wh-determiners compared to all tags, number of unique types of tags in the text)(2).

#### Paralinguistic features

Paralinguistic features were designed to capture changes in emotional state (like flattened affect) and articulation (proxy of changes in motor control). A unique, proprietary combination of 116 paralinguistic parameters was extracted from audio files, both at the level of task and in 10- second frames with 5 seconds overlap. Our feature set is comprised of temporal parameters (e.g. average syllable and pause ratio), prosodic parameters (e.g. glottal pulse period, jitter, shimmer), frequency parameters (e.g. F1 frequency mean), spectral parameters (e.g., Mel-frequency cepstral coefficients) and amplitude parameters (e.g., loudness).

#### Modelling Outcomes

Following the automated transcription and feature extraction, the speech and language features were used to address the stated objectives, as summarized in Figure 1. Data analyses were conducted in Python 3.7, Jupyter Notebook v6. 5.3.

**Figure 1:**
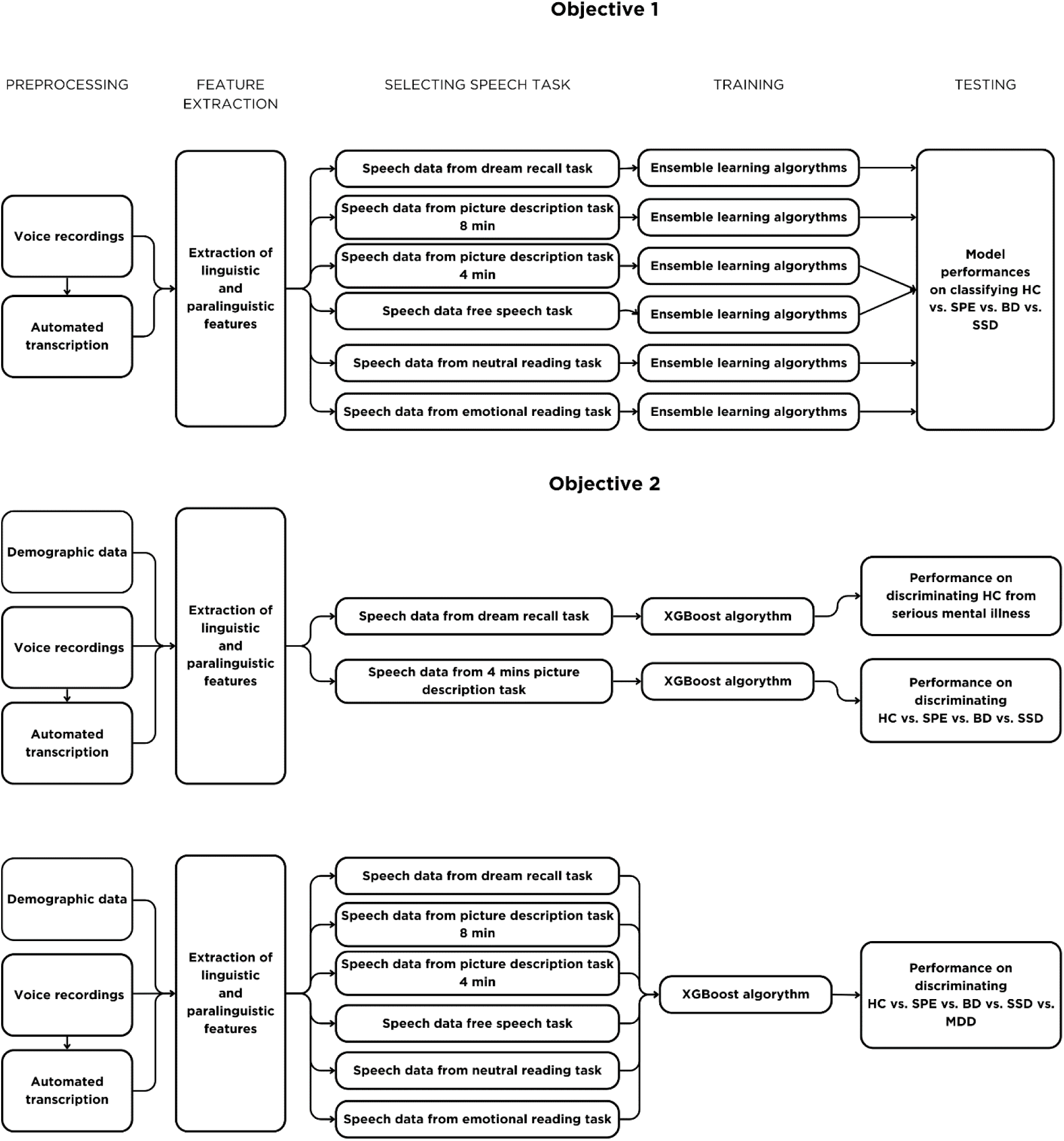
Overview of machine learning pipeline and modelling outcomes

**Figure 2:**
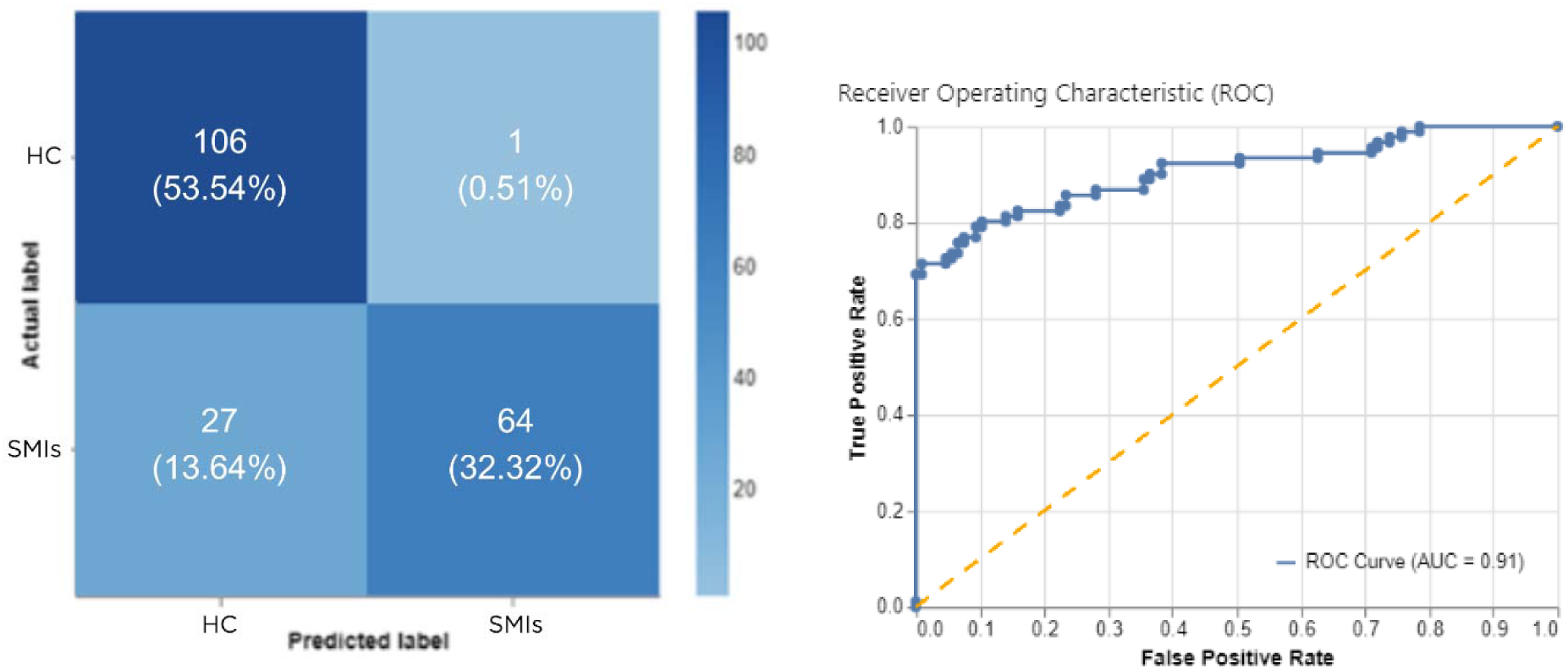
Confusion matrix (left) and ROC curve (Receiver Operating Characteristic curve) (right) of discriminating HC subjects and people with serious mental illness (SSD and BD) **Left:** Confusion matrix shows the true positive (TP)(right, down), true negative (TN)(left, up), false positive (FP) (up, right), and false negative (FN) (down, left) values on the test set. Shades reflect the number of cases in a given rubric. **Right** : ROC curve (Receiver Operating Characteristic curve) is a graphical representation of a classifier’s performance. It plots the true positive rate (sensitivity) against the false positive rate (1-specificity) at various thresholds, showing how well the model can distinguish between classes. The area under the ROC curve (AUC) quantifies the classifier’s overall performance - higher AUC values indicate better performance.

Performance metrics of accuracy and balanced accuracy were used to compare predictive power. We choose these metrics because accuracy is a standard, common metric used in ML applications that makes our results comparable with previous findings, and balanced accuracy accounts for the sensitivity of each class, making it a particularly useful metric when the dataset is imbalanced. Balanced accuracy prevents bias toward the majority class, providing a more realistic understanding of classification performance in our case. Accuracy is defined as the ratio of correctly predicted instances to the total number of instances in the dataset.

Mathematically, accuracy can be expressed as Accuracy = (TP + TN)/(TP + TN + FP +FN) * 100 where TP = True Positives, TN = True Negatives, FN = False Negatives, FP = False Positives. Balanced accuracy evaluates classification performance in multiclass problems by calculating the arithmetic mean of sensitivity (true positive rate) of each class. Sensitivity for each class is the number of true positive predictions for that class divided by the total number of actual instances of that class. The formula of balanced accuracy is Balanced Accuracy = (((TP/(TP+FN)+(TN/(TN+FP))) / 2where TP = True Positives, TN = True Negatives, FN = False Negatives, FP = False Positives.

#### Objective 1: Identify an optimal assessment approach for the online and remote collection of speech, in the context of assessing conditions on the psychosis spectrum

We first evaluated the predictive power of different types of speech tasks. We separated speech data per task and then fed each group of speech data corresponding to a given type of speech task into four types of ML algorithms (Random Forest, XGBoost, CatBoost, Extra Trees). Then, we compared the predictive power of the algorithms on the task of multiclass classification among the HC, SPE, BD, SSD groups. The four types of tested algorithms (Random Forest, XGBoost, CatBoost, Extra Trees) are types of ensemble learning, which combines the predictions of multiple individual models to create a stronger, more accurate, and robust model.

This approach has shown outstanding performance in speech and healthy applications, even when compared to more complex modelling approaches (e.g., Deep Neural Networks).

In this scenario, we aggregated data per file and per participant, taking the mean of different features at the level of the audio file and participant. ML algorithms were trained on 70% of the dataset using leave-one-out cross-validation and validated on the stratified, remaining 30% of the dataset.

#### Objective 2: Evaluate whether our ML pipeline can discriminate among different conditions on the schizophrenia-bipolar spectrum (SSD-BD-SPE), help-seeking comparison subjects (MDD), and healthy controls (HC) at varying layers of analysis

In the second part of our analysis, we wanted to test the discrimination ability of online collected speech at increasing levels of diagnostic complexity (Figure 1, Objective 2). Therefore firstly, we used the relatively simple task of binary classification when our speech-based machine learning pipeline had to discriminate between HC and serious mental illness (SSD and BD), modelling discrimination between relatively distinct conditions. Then, to increase complexity, we tested our speech-based ML pipeline in multiclass classification problem (4 classes) when it the model had to discriminate among different conditions on the psychosis spectrum (SPE, BD, SSD) and HC simultaneously. In addition to reducing the random chance of correct classification this task tested the model’s ability to discriminate between conditions that are less distinct, show overlapping symptom profile and challenging to separate using other biomarkers compared to the binary classification task. Lastly, we tested our speech-based ML pipeline in multiclass classification problem (5 classes) when the model also had to discriminate a help-seeking psychiatric controls (MDD) in addition to previous psychosis continuum classes, adding another layer of diagnostic complexity to the task.

We then tested whether our ML pipeline, based an optimised 5-minute speech battery, could discriminate between healthy controls and people with serious mental illness, defined as the combination of our SSD and BD groups). Basic demographic variables of age, gender and ethnicity were added to the pipeline as additional predictive features. Demographic and speech features were used to train an ensemble learning algorithm, XGBoost, as this algorithm overperformed other ensemble methods (Random Forest, CatBoost, Extra Trees) in the previous experiment. XGBoost was optimised for Area Under the ROC Curve (AUC), using leave-one-out cross-validation in the training set. A 70-30% train-test split ratio was applied to validate performance on the test set.

Next, we tested how our ML pipeline, based on the optimised 5-minutes speech battery, performed on the multiclass classification problem of discriminating between HC, SPE, BD and SSD groups. The same feature set was used, but we applied stratified SMOTE (Synthetic Minority Over-sampling Technique) on the training data to maximise the model’s training capabilities on the unbalanced dataset. The same ensemble learning algorithm, XGBoost, was applied on the feature set to train the model using leave-one-out crossvalidation. A 70-30% train-test split ratio was applied to validate performance on a stratified test set.

#### Evaluate whether our ML pipeline can further discriminate across the psychosis spectrum and major depressive disorder (HC-SPE-BD-SSD-MDD)

Finally, we evaluated whether our ML pipeline could discriminate across all 5 groups simultaneously: HC, SPE, SSD, BD, and MDD, where the latter is a purely affective condition. We designed this testing procedure as a proxy to evaluate whether our ML architecture is able to learn disorder-specific alterations, in contrast to discriminating between different severity and symptomology of the psychosis spectrum. Given our brief speech battery was optimised for psychosis spectrum, we used all 16- 20 minutes of speech samples available in order to evaluate predictive power of speech independent from the speech elicitation method. Like in previous steps, an ensemble learning algorithm, XGBoost, was trained and tested using speech and demographic features. 30-fold cross-validation was applied to measure performance.

## Results

### Sample

The sample consisted of 1140 individuals and 22,650 minutes of speech. The groups were imbalanced, due to the feasibility constraints of recruiting individuals with serious mental illness. The SSD group consisted of 84 participants, the BD group consisted of 227 participants, the SPE group consisted of 343 participants, the MDD group consisted of 156 participants and the HC group consisted of 330 participants. Demographic characteristics of the sample are displayed in Table 2.

**Table 2:**
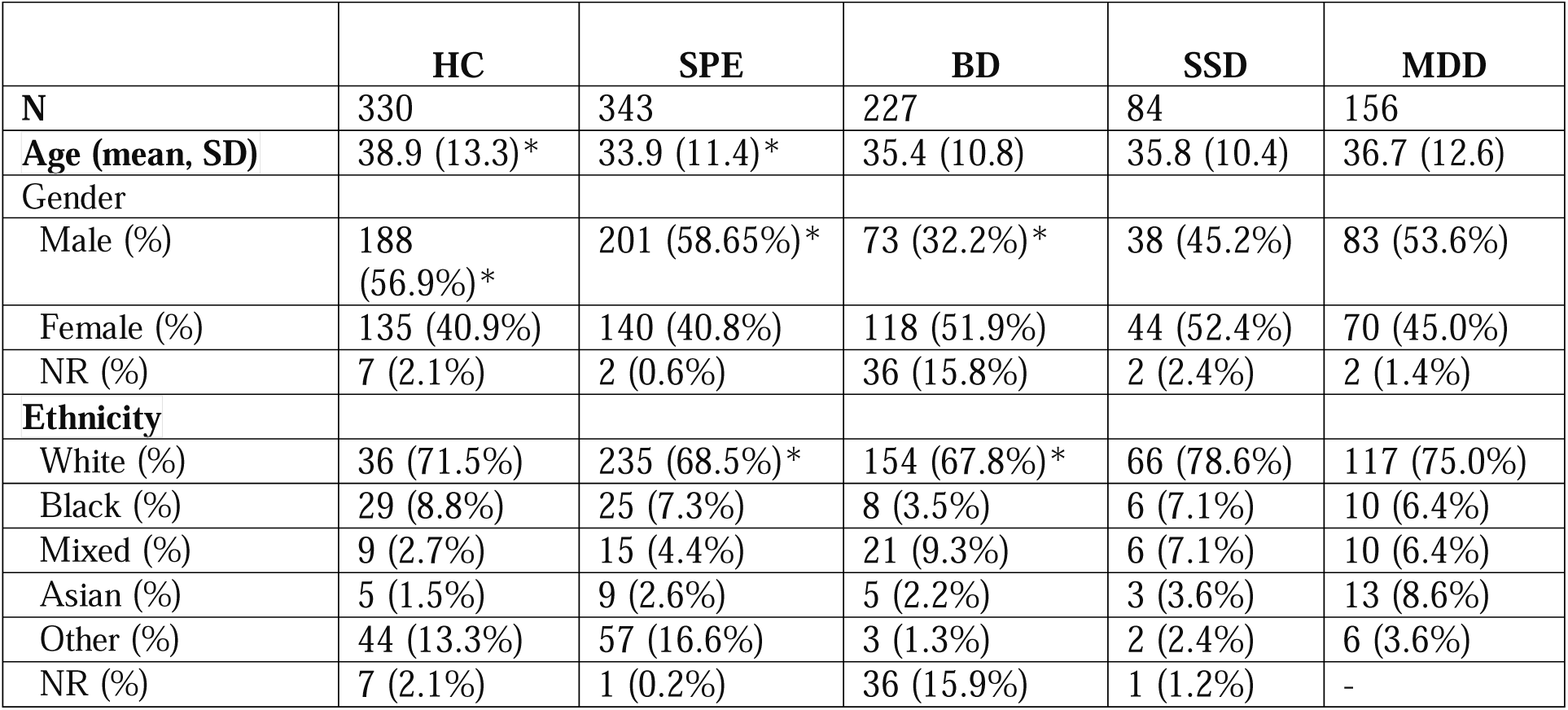
***Characteristics of the sample*** NR: Not reported *significant (p < 0.05) differences between groups according to Kruskal-Wallis and Dunn’s test

**Table 3:** Predictive power of different speech tasks on the classification of HC-SPE - BD-SSD groups Accuracy measures the overall correctness of a classification model. It is calculated as the ratio of correctly predicted instances to the total instances (Number of Correct Predictions/Total number of predictions). **Balanced accuracy:** a metric that takes into account imbalances in the class distribution. It calculates the average accuracy for each class and then averages those values. **Macro F1**: The F1 score is the harmonic mean of precision and recall. The macro F1 score calculates the F1 score for each class and then takes the average, giving equal weight to each class.

| Predictive power of different speech tasks on the classification of HC - SPE - BD - SSD groups |  |  |  |  |
| --- | --- | --- | --- | --- |
| Task type | Accuracy | Macro F1 | Balanced accuracy | Best model |
| Dream recall (~1 min) | 0.641 | 0.531 | 0.549 | XBoost |
| Picture description 8 pictures (~8 mins) | **0.669** | 0.567 | 0.577 | XBoost |
| Picture description 4 pictures (~4 mins) | 0.662 | **0.675** | **0.665** | XBoost |
| Free speech (~ 4 mins) | 0.623 | 0.63 | 0.627 | XBoost |
| Neutral text reading (~1 min) | 0.528 | 0.542 | 0.527 | XBoost |
| Emotional text reading (~ 3 mins) | 0.595 | 0.548 | 0.565 | XBoost |

### Comparing assessment approaches

Using all of the 20 minutes of speech recordings without demographic information, the predictive power of different tasks ranged between 52.8- 66.9% accuracy for discriminating among HC, SPE, BD and SSD groups (Table 4). Best performing models were consistently XBoost algorithms. The neutral text reading task had the lowest predictive power, and the picture description task had the highest. Sample duration did not show a strong connection with predictive power. For example, 8 minutes of picture description could achieve a slight improvement in performance compared to the dream recall task (1 minute), and in some metrics, underperformed the condition when we only considered the description of four pictures.

Considering time constraints and predictive power, we optimised our short speech task as 4 minutes of picture description and one dream recall task.

### Discriminating between healthy control and serious mental illness: HC vs. SSD/BD

Using computational features derived from the optimised 5 minutes of speech data and basic demographic information, our machine learning model could discriminate between HC group and individuals with serious mental illness (SSD and BD) with 86% accuracy (AUC: 0.91, Recall: 0.7, Precision: 0.98) in the test set.

### Discriminating among the schizophrenia-bipolar spectrum and healthy controls: HC vs. SPE vs. BD vs. SSD

Using the same, 5 minutes of speech from this optimised speech task and demographic variables, our machine learning model could discriminate between HC, SPE, BD and SSD groups simultaneously with 86% accuracy (F1 macro = 0.855, Recall Macro: 0.86, Precision Macro: 0.86) in our testing set.

### Discriminating among 5-classes: HC vs. MDD vs. SPE vs. BD vs. SSD

Using all of the speech data available (around 16 mins), our machine learning model could discriminate between HC, SPE, BD, SSD and MDD groups with 76% accuracy (F1 macro: 0.757, Recall Macro: 0.758, Precision Macro: 0.766) (Figure 3).

**Figure 3:**
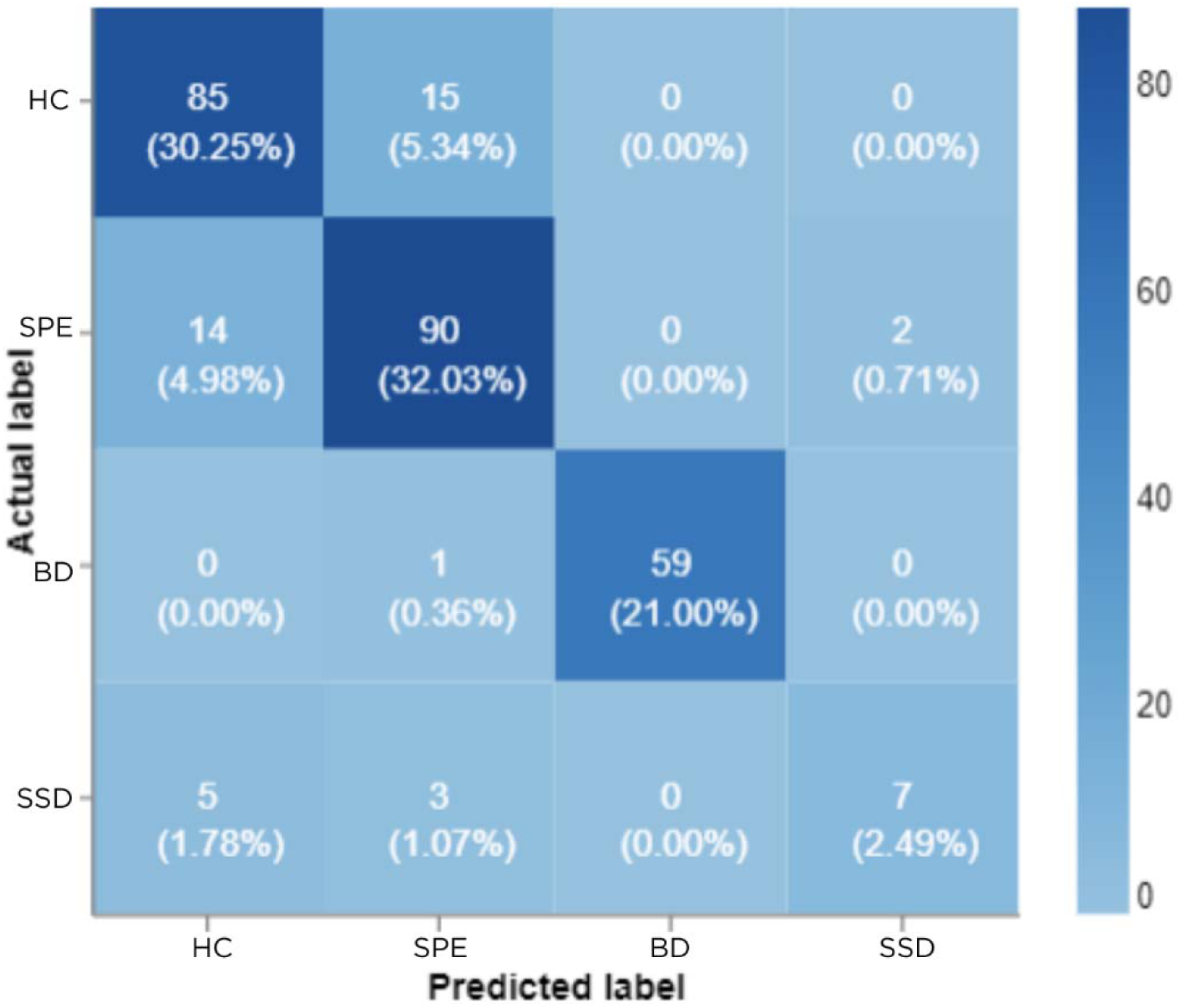
Confusion matrix shows the true positive (TP)(right, down), true negative (TN)(left, up), false positive (FP) (up, right), and false negative (FN) (down, left) values in the test set. Shades reflect the number of cases in a given rubric.

**Figure 4:**
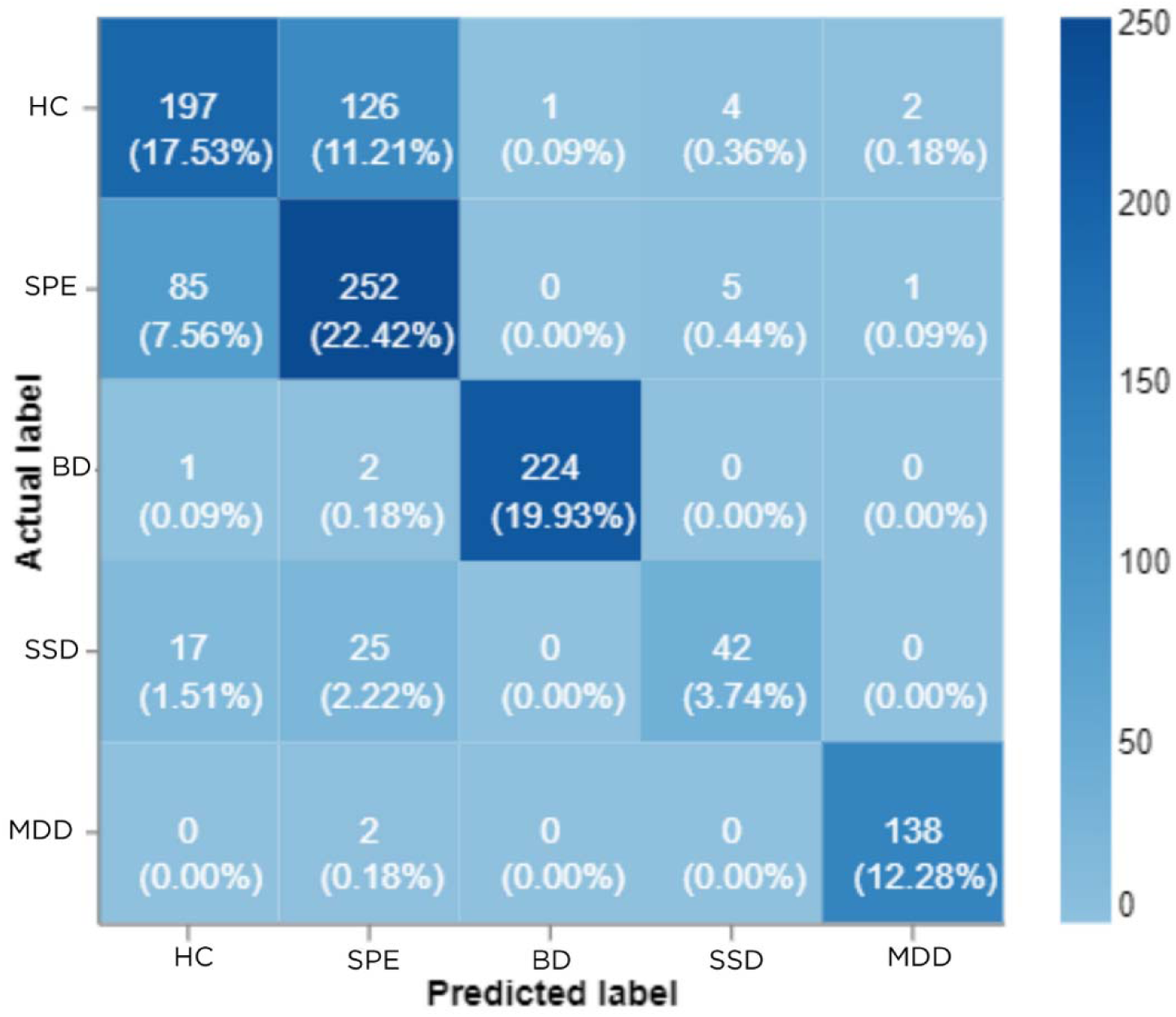
Confusion matrix shows the true positive (TP)(right, down), true negative (TN)(left, up), false positive (FP) (up, right), and false negative (FN) (down, left) values. Shades reflect the number of cases in a given rubric.

## Discussion

### Principal findings

In this study, we demonstrated the feasibility of using online-collected, self-recorded speech data to classify different conditions on the psychosis spectrum and MDD. The results demonstrate a fully automated, scalable approach that requires only 5 minutes of speech. More specifically, our findings highlight that speech, even when collected remotely and online, has a sufficient degree of between-group variability to discriminate between different forms and stages of psychosis spectrum conditions, as well as to discriminate between affective and psychotic conditions.

Our first objective was to compare the predictive power of different online speech-based tasks to discern different conditions on the psychosis spectrum. The results yielded showed that each task had different predictive power, and that increasing the length of a speech sample does not necessarily translate to better predictive power. Tasks which included the generation of speech – rather than the recitation of prewritten text - were more informative. We speculate that this observation might stem from the additional variation in the textual information yielded by the response-based tasks, relative to reading tasks that only generate variance in paralinguistic parameters. While affective changes and motor control deviations can be clearly captured from acoustic information, thought disorder, one of the key symptoms of psychotic conditions, is more connected to language alterations (22,50,51,53–57) It’s reasonable then that the tasks leading to variance in language suitable for capturing these conditions. Our findings are consistent with previous research that showed the suitability of a dream recall task(58) and picture description tasks(47), particularly when compared against interview settings of wakening reports.

Once we identified the optimal speech task battery to utilise, we then trained and tested our machine learning algorithms to assess its classification efficacy. Our ML pipeline showed excellent performance in a classification task between HC and individuals with serious mental illness. Previous studies leveraging either acoustic or NLP-based models demonstrated varied performance ranging from moderate (min. AUC = 0.63) to excellent (max accuracy = 97.68%) in discriminating between healthy controls and patients with schizophrenia (17,51,59–65). Prior reports also have explored the use of AI techniques to discriminate between HC and patients with BD, with moderate performance reported (AUC ranges between 0.742 and 0.8)(66,67). It is difficult to directly compare the performance of our ML pipelines, given the differences in our study designs. Previous work used more homogeneous samples, had smaller sample sizes (N raged between 15-288) and generally collated longer speech samples in offline, laboratory conditions. We had a diverse, large sample and utilised online, remote speech data collection. Yet, the performance of our model (AUC = 0.91) was comparable and possibly improved upon the best-performing models in the field. Broadly, our results suggest that strong performance and accuracy in identifying psychotic conditions can be maintained even when using scalable assessment methods in a larger and more robust sample.

In addition to this binary classification task, our ML pipeline showed relatively strong performance in classifying across HC, SPE (individuals with significant psychosis symptoms but not meeting threshold for a clinical psychotic disorder diagnosis), BD, and SSD groups. Notably, the overall accuracy of the model did not drop when we introduced two extra classes into the model compared to the binary classification (HCvs. SSD/MD), suggesting that speech has sufficient variance between different schizophrenia-bipolar spectrum conditions to enable high discriminatory power.

To our knowledge, this is the first time that AI-based algorithms have been used for diagnostic classification across this range of disorders – no prior study has leveraged linguistic markers to evaluate classification across more than 3 conditions on the psychotic spectrum. The only comparable study is from Espinola et al (68) who trained a model to discriminate between major depression, bipolar disorder, schizophrenia, anxiety and healthy control, using paralinguistic parameters from speech samples recorded offline. They achieved similar performance (accuracy = 76%) on a small sample of 76 people. Our results reinforce and expand on this work, using a more robust, short sample of self-recorded, online collected speech. Spencer et al(22) have also experimented with discriminating healthy controls, people at clinical high risk and first-episode patients with automated language markers. They reported associations of different speech markers with specific conditions and transition to psychosis from at-risk state, however, their samples were limited in size, and they did not apply predictive modelling which makes it difficult to compare results. Studies that aimed to discriminate between bipolar disorder and schizophrenia, reported high performance (AUC ranged between 0.87-0.92)(66,69).

While these results are promising, some misclassifications did also occur. Notably, most misclassification occurred between the SPE and HC groups. We hypothesise that the reason underlying this is the relatively weak proxy of the SPE group labelled in our study. Caseness for this SPE group was defined as having endorsed more than 6 symptoms on the PQ-16 scale.

Although this threshold has previously been suggested as a screening threshold for clinical at-risk services (43), it is known to provide sensitivity at the cost of poor specificity. Consequently, it is reasonable to presume that a good portion of this SPE group does not meet the threshold for clinical high risk of psychosis, and overlaps with a “healthy” individual. It is likely that this broad definition of psychosis spectrum status might have blurred the threshold between our healthy group and the SPE group. We speculate that using a more stringent threshold may reduce the number of misclassifications, but that is beyond the scope of this investigation.

There was also some misclassification between healthy individuals and the SSD group. This may suggest that our definition of healthy control subjects was also relatively lenient. Individuals were considered healthy if they did not report a clinical diagnosis of mental disorders, and were below the threshold on the PQ-16 and PHQ-8; however, evidently, these criteria do not necessarily imply that these individuals do not suffer from mental health conditions. Along similar lines, we speculate that the majority of the participants in the SSD group were stable and in remission while taking part in the study. This is a fair assumption given the proactivity, concentration, online presence and continuous engagement required to comply with the remote study. Although the composition of the group allows us to assume that the model captures trait-related changes in speech as opposed to disorder state-related changes, it also decreases the observable differences between the speech of the HC group and the SSD group. Therefore, future studies should focus on validating and replicating current findings on clinically validated and labelled data. Also, future research is needed to validate the performance of automated, speech-based AI models to captures changes in dynamic, state-like variations like symptom severity.

This misclassification pattern was replicated in our final analyses, where we trained and tested our algorithms to discriminate across all five groups, including MDD. This reinforces the notion that the definition of these groups were a relatively lenient proxy. Nevertheless, the best-performing ML models still achieved acceptable performance on the discrimination task.

Notably, the model performed well in discerning between serious mental illness, HC and MDD, as well as between MDD and BD. Differential diagnosis across these conditions is a challenge in clinical settings, particularly in primary care settings (60,70). As such, the capability of our ML pipeline to distinguish between these conditions suggests that there is a strong potential translational benefit of our technology into clinical care, complementing clinical decision-making.

## Strengths & Limitations

This study possesses several strengths. We included multiple psychiatric conditions across the psychosis and affective spectrum, rather than focusing solely on binary algorithmic classifications. Furthermore, our sample is, to our knowledge, the largest to date, reinforcing the robustness of our findings across a heterogeneous population. As study participants were not recruited during the acute stage of their illness, our results are more likely to rely on trait variations in speech specific to each disorder rather than “state” related alterations. In addition, the online nature of all data collection and assessments improves the ecological validity of our findings, reflective of real-world situations.

It is also important to acknowledge the limitations of this study. Firstly, our study does not comprehensively consider cognitive measures, which could potentially confound our results. Cognition has previously been connected to speech changes and is a predictor of psychosis severity (71). However, the magnitude of cognition’s effects is ambiguous: some studies suggest that cognition has a strong effect on some features (especially connectivity-based ones), though it appears independent of other features (52,55,71–73). While we included several demographic variables in our analyses, it was beyond the scope to consider cognitive-related measures. Future research should consider certain cognitive variables and their impact on the predictive power of speech-based models; for example, education can be used as a proxy.

Secondly, this study utilizes self-report questionnaires and self-reports of clinical diagnosis as a proxy for having a certain condition. This is particularly pertinent to our definition of the SPE, which was defined using the self-report PQ-16 questionnaire. Though we employed a previously used cut-off, the reliance on self-reports might have introduced biases into our analyses. It is feasible that more stringent or strictly clinical labels would improve the performance of our models, given this would reduce the potential overlaps between classes. It is therefore imperative to further validate in additional clinical samples.

Thirdly, it is important to underscore the unbalanced nature of our sample (e.g. SSD groups was underrepresented compared to HC or SPE), which might well skew results, particularly when comparing different conditions. However, we employed analytical methods (specifically SMOTE, comparing balanced accuracy, and macro F1 scores) to mitigate the effects of this unbalanced sample.

Fourthly, the current research was designed as a proof-of-concept study for a fully automated, remote, short, speech-based assessment of different conditions on the psychosis spectrum.

Accordingly, we kept our ML models lean and comparable to each other. However, it is likely that deploying more complex ML architectures (such as Deep Neural Networks) and experimenting with a wider range of models and hyperparameter optimisation of such models would lead to better performance. Therefore, we interpret the reported model capabilities as promising findings rather than an upper marker of the potential capabilities of fully automated, remote, short, speech-based assessment technologies.

Fifthly, the current study had certain exclusion criteria that may restrict our findings’ generalisability. The sample only included native English speakers and excluded people with neurodiversity. Further research should be conducted in samples that include neurodiverse groups and non-English languages, to evaluate whether our models are as effective in more representative samples, a crucial step to application in clinical setting.

Finally, the cross-sectional nature of our study restricts our ability to draw conclusions about causality and long-term trends. Longitudinal studies would help address this gap, allowing researchers to train models for the prediction of symptom changes, and allowing for the continuous emergence of speech alterations. The use of remote, quick assessments, as demonstrated in our study, might help facilitate such longitudinal studies, by enabling a more convenient way to continuously collect speech samples.

## Conclusions

This study provides valuable insights into the scalability of speech-based assessment methods for the identification and classification of psychosis spectrum and affective conditions. Our findings broadly suggest that our ML pipeline learnt disorder- and severity-specific information from speech. We leveraged a large and robust sample to demonstrate best-in-class classification accuracy across psychosis spectrum conditions, using online, self-recorded speech data. Furthermore, this is the first report of accurate classification across 5 psychiatric groups. Overall, this study showcases the huge potential of online, remote-collected speech for application in clinical contexts.

## Funding

This research has been funded by Innovate UK, Fast Start Grant. SXT is supported by the NIH (K23 MH130750) and the Brain and Behavior Research Foundation Young Investigator Grant.

## Authors’ contribution

J. O., and S. X. T. conceptualised the study. E. W., R. Ch., J. O. administered the project and conducted data collection.J.O. wrote the first version of the manuscript. J.O. and O. M. analysed the data and ran experiments. All authors contributed to the investigation, interpretation, review and editing of the manuscript. All authors read and approved the final version of the manuscript.

## Conflict of interest

J.O, E.W., R. Ch are co-founders and employees of Psyrin and owns equity for Psyrin. SXT owns equity and serves as a consultant for North Shore Therapeutics, received research funding and serves as a consultant for Winterlight Labs, and is on the advisory board and owns equity for Psyrin.

## Data Availability

All data produced in the present study are available upon reasonable request to the authors for IRB-approved research purposes.

## Notes

### Author Declarations

Ethics committee of Advarra Inc. (IRB Registration Number: 00000971) waived ethical approval for this work.

## References

(1) Corcoran CM, Mittal VA, Bearden CE, E. Gur R, Hitczenko K, Bilgrami Z, et al. Language as a biomarker for psychosis: A natural language processing approach. Schizophrenia research 2020 Dec;226:158–166.

(2) Corcoran CM, Cecchi GA. Using Language Processing and Speech Analysis for the Identification of Psychosis and Other Disorders. Biological psychiatry : cognitive neuroscience and neuroimaging 2020 Aug;5(8):770–779.

(3) Palaniyappan L. More than a biomarker: could language be a biosocial marker of psychosis? NPJ schizophrenia 2021 Aug 31,;7(1):42.

(4) Parola A, Simonsen A, Lin JM, Zhou Y, Wang H, Ubukata S, et al. Voice Patterns as Markers of Schizophrenia: Building a Cumulative Generalizable Approach Via a Cross-Linguistic and Meta-analysis Based Investigation. Schizophrenia bulletin 2023 Mar 22,;49(Suppl_2):S125- S141.

(5) Schneider K, Leinweber K, Jamalabadi H, Teutenberg L, Brosch K, Pfarr J, et al. Syntactic complexity and diversity of spontaneous speech production in schizophrenia spectrum and major depressive disorders. NPJ schizophrenia 2023 May 29,;9(1):35.

(6) Tang SX, Kriz R, Cho S, Park SJ, Harowitz J, Gur RE, et al. Natural language processing methods are sensitive to sub-clinical linguistic differences in schizophrenia spectrum disorders. NPJ schizophrenia 2021 May 14,;7(1):25.

(7) Tognin S, van Hell HH, Merritt K, Winter-van Rossum I, Bossong MG, Kempton MJ, et al. Towards Precision Medicine in Psychosis: Benefits and Challenges of Multimodal Multicenter Studies—PSYSCAN: Translating Neuroimaging Findings From Research into Clinical Practice. Schizophrenia bulletin 2020 Feb 26,;46(2):432-441.

(8) Gupta T, Hespos SJ, Horton WS, Mittal VA. Automated analysis of written narratives reveals abnormalities in referential cohesion in youth at ultra high risk for psychosis. Schizophrenia research 2017;192:82–88.

(9) Corcoran CM, Carrillo F, Fernández-Slezak D, Bedi G, Klim C, Javitt DC, et al. Prediction of psychosis across protocols and risk cohorts using automated language analysis. World psychiatry 2018 Feb;17(1):67–75.

(10) Rezaii N, Walker E, Wolff P. A machine learning approach to predicting psychosis using semantic density and latent content analysis. NPJ Schizophrenia 2019 Jun 13,;5(1):9-12.

(11) Nicholas Cummins, Judith Dineley, Pauline Conde, Faith Matcham, Sara Siddi, Femke Lamers, et al. Multilingual markers of depression in remotely collected speech samples. npj Digital Medicine 2022.

(12) Hoffman GMA, Gonze JC, Mendlewicz J. Speech Pause Time as a Method for the Evaluation of Psychomotor Retardation in Depressive Illness. British journal of psychiatry 1985 May;146(5):535–538.

(13) Voppel A, de Boer J, Brederoo S, Schnack H, Sommer I. Quantified language connectedness in schizophrenia-spectrum disorders. Psychiatry research 2021 Oct;304:114130.

(14) Chang X, Zhao W, Kang J, Xiang S, Xie C, Corona-Hernández H, et al. Language abnormalities in schizophrenia: binding core symptoms through contemporary empirical evidence. NPJ schizophrenia 2022 Nov 12,;8(1):95.

(15) Alonso-Sánchez MF, Ford SD, MacKinley M, Silva A, Limongi R, Palaniyappan L. Progressive changes in descriptive discourse in First Episode Schizophrenia: a longitudinal computational semantics study. NPJ schizophrenia 2022 Apr 12,;8(1):36.

(16) Mackinley M, Chan J, Ke H, Dempster K, Palaniyappan L. Linguistic determinants of formal thought disorder in first episode psychosis. Early intervention in psychiatry 2021 Apr;15(2):344–351.

(17) De Boer JN, Voppel AE, Brederoo SG, Schnack HG, Truong KP, Wijnen FNK, et al. Acoustic speech markers for schizophrenia-spectrum disorders: A diagnostic and symptom-recognition tool. Psychological medicine 2021 Aug 4,:1–11.

(18) Ciampelli S, de Boer JN, Voppel AE, Corona Hernandez H, Brederoo SG, van Dellen E, et al. Syntactic Network Analysis in Schizophrenia-Spectrum Disorders. Schizophrenia bulletin 2023 Mar 22,;49(Suppl_2):S172-S182.

(19) Palaniyappan L, Mota NB, Oowise S, Balain V, Copelli M, Ribeiro S, et al. Speech structure links the neural and socio-behavioural correlates of psychotic disorders. Progress in neuro- psychopharmacology & biological psychiatry 2019 Jan 10,;88:112–120.

(20) Corona Hernández H, Corcoran C, Achim AM, de Boer JN, Boerma T, Brederoo SG, et al. Natural Language Processing Markers for Psychosis and Other Psychiatric Disorders: Emerging Themes and Research Agenda From a Cross-Linguistic Workshop. Schizophrenia bulletin 2023 Mar 22,;49(Suppl_2):S86-S92.

(21) Tang SX, Kriz R, Cho S, Park SJ, Harowitz J, Gur RE, et al. Natural language processing methods are sensitive to sub-clinical linguistic differences in schizophrenia spectrum disorders. NPJ schizophrenia 2021 May 14,;7(1):25.

(22) Spencer TJ, Thompson B, Oliver D, Diederen K, Demjaha A, Weinstein S, et al. Lower speech connectedness linked to incidence of psychosis in people at clinical high risk. Schizophrenia research 2021 Feb;228:493–501.

(23) Bedi G, Carrillo F, Cecchi GA, Slezak DF, Sigman M, Mota NB, et al. Automated analysis of free speech predicts psychosis onset in high-risk youths. NPJ schizophrenia 2015;1(1):15030.

(24) Bedwell J, Cohen A, Trachik B, Deptula A, Mitchell J. Speech Prosody Abnormalities and Specific Dimensional Schizotypy Features: Are Relationships Limited to Male Participants? The journal of nervous and mental disease 2014 Oct;202(10):745–751.

(25) Cohen AS, Lee Hong S. Understanding constricted affect in schizotypy through computerized prosodic analysis. Journal of personality disorders 2011;25(4):478–491.

(26) Cohen AS, Cox CR, Cowan T, Masucci MD, Le TP, Docherty AR, et al. High Predictive Accuracy of Negative Schizotypy With Acoustic Measures. Clinical psychological science 2022 Mar;10(2):310–323.

(27) Cohen A, Elvevåg B. Automated computerized analysis of speech in psychiatric disorders. Current opinion in psychiatry 2014 May;27(3):203–209.

(28) Hinzen W, Rosselló J, McKenna P. Can delusions be understood linguistically? Cognitive neuropsychiatry 2016 Jul 3,;21(4):281-299.

(29) Hinzen W, Rosselló J. The linguistics of schizophrenia: thought disturbance as language pathology across positive symptoms. Frontiers in psychology 2015 Jul 16,;6:971.

(30) Stein F, Gruber M, Mauritz M, Brosch K, Pfarr J, Ringwald KG, et al. Brain Structural Network Connectivity of Formal Thought Disorder Dimensions in Affective and Psychotic Disorders. Biological psychiatry (1969) 2023 May 18,.

(31) Ehlen F, Montag C, Leopold K, Heinz A. Linguistic findings in persons with schizophrenia-a review of the current literature. Frontiers in psychology 2023;14:1287706.

(32) Abi-Dargham A, Moeller SJ, Ali F, DeLorenzo C, Domschke K, Horga G, et al. Candidate biomarkers in psychiatric disorders: state of the field. World psychiatry 2023 Jun;22(2):236–262.

(33) Analyzing acoustic and prosodic fluctuations in free speech to predict psychosis onset in high-risk youths. : IEEE; Jul 2020.

(34) Birnbaum ML, Abrami A, Heisig S, Ali A, Arenare E, Agurto C, et al. Acoustic and Facial Features From Clinical Interviews for Machine Learning-Based Psychiatric Diagnosis: Algorithm Development. JMIR mental health 2022 Jan 24,;9(1):e24699.

(35) Keshavan MS, Morris DW, Sweeney JA, Pearlson G, Thaker G, Seidman LJ, et al. A dimensional approach to the psychosis spectrum between bipolar disorder and schizophrenia: The Schizo-Bipolar Scale. Schizophrenia research 2011 Dec 1,;133(1):250-254.

(36) Jablensky A. The diagnostic concept of schizophrenia: its history, evolution, and future prospects. Dialogues in clinical neuroscience 2010;12(3):271–287.

(37) Tamminga CA, Pearlson G, Keshavan M, Sweeney J, Clementz B, Thaker G. Bipolar and Schizophrenia Network for Intermediate Phenotypes: Outcomes Across the Psychosis Continuum. Schizophrenia bulletin 2014 Mar 1,;40(Suppl 2):S131-S137.

(38) Reininghaus U, Böhnke JR, Chavez-Baldini U, Gibbons R, Ivleva E, Clementz BA, et al. Transdiagnostic dimensions of psychosis in the Bipolar-Schizophrenia Network on Intermediate Phenotypes (B-SNIP). World psychiatry 2019 Feb;18(1):67–76.

(39) Schneider K, Leinweber K, Jamalabadi H, Teutenberg L, Brosch K, Pfarr J, et al. Syntactic complexity and diversity of spontaneous speech production in schizophrenia spectrum and major depressive disorders. NPJ schizophrenia 2023 May 29,;9(1):35.

(40) Berardi M, Brosch K, Pfarr J, Schneider K, Sültmann A, Thomas-Odenthal F, et al. Relative importance of speech and voice features in the classification of schizophrenia and depression. Translational psychiatry 2023 Sep 19,;13(1):298.

(41) Koutsouleris N, Meisenzahl EM, Borgwardt S, Riecher-Rössler A, Frodl T, Kambeitz J, et al. Individualized differential diagnosis of schizophrenia and mood disorders using neuroanatomical biomarkers. Brain (London, England : 1878) 2015 Jul 1,;138(Pt 7):2059-2073.

(42) Ising HK, Veling W, Loewy RL, Rietveld MW, Rietdijk J, gt S, et al. The Validity of the 16- Item Version of the Prodromal Questionnaire (PQ-16) to Screen for Ultra High Risk of Developing Psychosis in the General Help-Seeking Population. Schizophrenia bulletin 2012 Nov 1,;38(6):1288-1296.

(43) McDonald M, Christoforidou E, Van Rijsbergen N, Gajwani R, Gross J, Gumley AI, et al. Using Online Screening in the General Population to Detect Participants at Clinical High-Risk for Psychosis. Schizophrenia bulletin 2019 Apr 25,;45(3):600-609.

(44) Rancans E, Vrublevska J, Trapencieris M, Snikere S, Ivanovs R, Logins R, et al. Validity of patient health questionnaire (PHQ-9) in detecting depression in primary care settings in Latvia – the results of the National Research Project BIOMEDICINE. European neuropsychopharmacology 2016;26:S481.

(45) Kroenke K, Spitzer RL. The PHQ-9: A New Depression Diagnostic and Severity Measure. Psychiatric annals 2002 Sep 1,;32(9):509-515.

(46) Kroenke K, Strine TW, Spitzer RL, Williams JBW, Berry JT, Mokdad AH. The PHQ-8 as a measure of current depression in the general population. Journal of affective disorders 2009 Apr 1,;114(1):163-173.

(47) Morgan SE, Diederen K, Vértes PE, Ip SHY, Wang B, Thompson B, et al. Natural Language Processing markers in first episode psychosis and people at clinical high-risk. Translational psychiatry 2021 Dec 13,;11(1):630.

(48) Olah J, Diederen K, Gibbs-Dean T, Kempton MJ, Dobson R, Spencer T, et al. Online speech assessment of the psychotic spectrum: Exploring the relationship between overlapping acoustic markers of schizotypy, depression and anxiety. Schizophrenia research 2023 Apr 18,.

(49) Mikolov T, Chen K, Corrado G, Dean J. Efficient estimation of word representations in vector space. arXiv.org 2013 Jan 16,.

(50) Nettekoven CR, Diederen K, Giles O, Duncan H, Stenson I, Olah J, et al. Semantic Speech Networks Linked to Formal Thought Disorder in Early Psychosis. Schizophrenia bulletin 2023 Mar 22,;49(Suppl_2):S142-S152.

(51) Mota NB, Copelli M, Ribeiro S. Thought disorder measured as random speech structure classifies negative symptoms and schizophrenia diagnosis 6 months in advance. NPJ Schizophrenia 2017;3(1):18–10.

(52) Mota NB, Weissheimer J, Finger I, Ribeiro M, Malcorra B, Hübner L. Speech as a Graph: Developmental Perspectives on the Organization of Spoken Language. Biological psychiatry : cognitive neuroscience and neuroimaging 2023 Apr 19,.

(53) Premkumar P, Kuipers E, Kumari V. The path from schizotypy to depression and aggression and the role of family stress. European psychiatry 2020 Jul 30,;63(1):e79.

(54) Xu W, Wang W, Portanova J, Chander A, Campbell A, Pakhomov S, et al. Fully automated detection of formal thought disorder with Time-series Augmented Representations for Detection of Incoherent Speech (TARDIS). Journal of biomedical informatics 2022 Feb;126:103998.

(55) Mota NB, Vasconcelos NAP, Lemos N, Pieretti AC, Kinouchi O, Cecchi GA, et al. Speech Graphs Provide a Quantitative Measure of Thought Disorder in Psychosis. PLoS ONE 2012 Apr 9,;7(4):e34928.

(56) Harvey PD. Speech competence in manic and schizophrenic psychoses: The association between clinically rated thought disorder and cohesion and reference performance. Journal of abnormal psychology (1965) 1983 Aug;92(3):368-377.

(57) Ayer A, Yalınçetin B, Aydınlı E, Sevilmiş Ş, Ulaş H, Binbay T, et al. Formal Thought Disorder in First-Episode Psychosis. Comprehensive Psychiatry 2016 Oct 1,;70:209–215.

(58) Mota NB, Furtado R, Maia PPC, Copelli M, Ribeiro S. Graph analysis of dream reports is especially informative about psychosis. Scientific Reports 2014 Jan 15,;4(1):3691.

(59) Automatic detection of incoherent speech for diagnosing schizophrenia. : Association for Computational Linguistics; Jan 1, 2018.

(60) Benoit J, Onyeaka H, Keshavan M, Torous J. Systematic Review of Digital Phenotyping and Machine Learning in Psychosis Spectrum Illnesses. Harvard review of psychiatry 2020 Sep;28(5):296–304.

(61) Martínez-Sánchez F, Muela-Martínez JA, Cortés-Soto P, García Meilán JJ, Vera Ferrándiz JA, Egea Caparrós A, et al. Can the Acoustic Analysis of Expressive Prosody Discriminate Schizophrenia? The Spanish journal of psychology 2015;18:E86.

(62) Huang Y, Lin Y, Liu C, Lee L, Hung S, Lo J, et al. Assessing Schizophrenia Patients Through Linguistic and Acoustic Features Using Deep Learning Techniques. TNSRE 2022;30:947–956.

(63) Silva AM, Limongi R, MacKinley M, Ford SD, Alonso-Sánchez MF, Palaniyappan L. Syntactic complexity of spoken language in the diagnosis of schizophrenia: A probabilistic Bayes network model. Schizophrenia research 2022 Jun 22,.

(64) Turek A, Machalska K, Chrobak AA, Tereszko A, Siwek M, Dudek D. Speech graph analysis of verbal fluency tests distinguish between patients with schizophrenia and healthy controls. European neuropsychopharmacology 2017 Oct;27:S914–S915.

(65) de Boer JN, Voppel AE, Brederoo SG, Schnack HG, Truong KP, Wijnen FNK, et al. Acoustic speech markers for schizophrenia-spectrum disorders: a diagnostic and symptom-recognition tool. Psychological medicine 2023 Mar 1,;53(4):1302-1312.

(66) Pan W, Deng F, Wang X, Hang B, Zhou W, Zhu T. Exploring the ability of vocal biomarkers in distinguishing depression from bipolar disorder, schizophrenia, and healthy controls. Front Psychiatry 2023 Jul 20;14:1079448.

(67) Naderi H, Behrouz Haji Soleimani, Matwin S. Multimodal Deep Learning for Mental Disorders Prediction from Audio Speech Samples. arXiv (Cornell University) 2020 Apr 13,.

(68) Wanderley Espinola C, Gomes JC, Mônica Silva Pereira J, dos Santos WP. Detection of major depressive disorder, bipolar disorder, schizophrenia and generalized anxiety disorder using vocal acoustic analysis and machine learning: an exploratory study. Res Biomed Eng 2022 Sep 1,;38(3):813-829.

(69) Making a distinction between schizophrenia and bipolar disorder based on temporal parameters in spontaneous speech. ; 2020.

(70) Buckley PF, Miller BJ, Lehrer DS, Castle DJ. Psychiatric Comorbidities and Schizophrenia. Schizophrenia bulletin 2009 Mar;35(2):383–402.

(71) Moberget T, Ivry RB. Prediction, Psychosis, and the Cerebellum. Biological psychiatry : cognitive neuroscience and neuroimaging 2019 Sep 1,;4(9):820–831.

(72) Mota NB, Weissheimer J, Finger I, Ribeiro M, Malcorra B, Hübner L. Speech as a Graph: Developmental Perspectives on the Organization of Spoken Language. Biological psychiatry : cognitive neuroscience and neuroimaging 2023 Apr 20,.

(73) Mota NB, Sigman M, Cecchi G, Copelli M, Ribeiro S. The maturation of speech structure in psychosis is resistant to formal education. NPJ Schizophrenia 2018 Dec 7,;4(1):25-10.

